# Optimized BERT-based NLP outperforms Zero-Shot Methods for Automated Symptom Detection in Clinical Practice

**DOI:** 10.1101/2025.04.21.25326037

**Authors:** Juan G. Diaz Ochoa, Natalie Layer, Jonas Mahr, Faizan E Mustafa, Christian U. Menzel, Martina Müller, Tobias Schilling, Gerald Illerhaus, Markus Knott, Alexander Krohn

## Abstract

**Background:** Large Language Nodels (LLMs) have raised broad expectations for clinical use, particularly in the processing of complex medical narratives. However, in practice, more targeted Natural Language Processing (NLP) approaches may offer higher precision and feasibility for symptom extraction from real-world clinical texts. NLP provides promising tools for extracting clinical information from unstructured medical narratives. However, few studies have focused on integrating symptom information from free texts in German, particularly for complex patient groups such as emergency department (ED) patients. The ED setting presents specific challenges: high documentation pressure, heterogeneous language styles, and the need for secure, locally deployable models due to strict data protection regulations. Furthermore, German remains a low-resource language in clinical NLP.

**Methods:** We implemented and compared two models for zero-shot learning—GLiNER and Mistral—and a fine-tuned BERT-based SCAI-BIO/BioGottBERT model for named entity recognition (NER) of symptoms, anatomical terms, and negations in German ED anamnesis texts in an on-premises environment in a hospital. Manual annotations of 150 narratives were used for model validation. The postprocessing steps included confidence-based filtering, negation exclusion, symptom standardization, and integration with structured oncology registry data. All computations were performed on local hospital servers in an on-premises implementation to ensure full data protection compliance.

**Results:** The fine-tuned SCAI-BIO/BioGottBERT model outperformed both zero-shot approaches, achieving an F1 score of 0.84 for symptom extraction and demonstrating superior performance in negation detection. The validated pipeline enabled systematic extraction of affirmed symptoms from ED-free text, transforming them into structured data. This method allows large-scale analysis of symptom profiles across patient populations and serves as a technical foundation for symptom-based clustering and subgroup analysis.

**Conclusions:** Our study demonstrates that modern NLP methods can reliably extract clinical symptoms from German ED free text, even under strict data protection constraints and with limited training resources. Fine-tuned models offer a precise and practical solution for integrating unstructured narratives into clinical decision-making. This work lays the methodological foundation for a new way of systematically analyzing large patient cohorts on the basis of free-text data. Beyond symptoms, this approach can be extended to extracting diagnoses, procedures, or other clinically relevant entities. Building upon this framework, we apply network-based clustering methods (in a subsequent study) to identify clinically meaningful patient subgroups and explore sex- and age-specific patterns in symptom expression.

## 1 Introduction

Clinical notes are valuable tools for describing patients’ phenomenal experiences, such as symptoms. These experiences, or *qualia*, refer to the subjective, qualitative aspects of perception that may not always directly reflect disease processes (Robinson 2023). In medical practice, physicians interpret these subjective reports and translate them into structured descriptions that guide diagnosis and treatment. This interpretive process is central to clinical reasoning, as physicians’ impressions and *gestalt* influence how symptoms are prioritized and linked to potential diagnoses (Tybjerg 2023). While standardized codes in electronic health records (EHRs), such as the ICD-10 or CEDIS, capture essential information, they fail to reflect the full complexity of patients’ lived experience, underscoring the importance of free-text clinical documentation in patient care.

Analyzing these narratives is therefore essential for a comprehensive understanding of the clinical context. However, anamnesis texts are typically unstructured, manually written, and highly variable in style (Raghavan et al. 2014). Although they contain rich information relevant for diagnosis, decision-making, and research, their unstructured nature hinders their systematic use. Hence, methods are needed to transform these texts into structured, analyzable formats.

While conventional machine learning models process structured data efficiently, they often fall short in reproducing the nuanced, contextual understanding of experienced physicians in complex scenarios (Kim et al. 2025). To be clinically applicable, models must preserve the semantic richness of clinical narratives rather than flattening them into overly simplified representations.

NLP technologies have already been used to extract structured elements from EHRs (Munzone et al. 2024), often employing transformer-based architectures such as BERT and tagging schemes such as BIO for named entity recognition (NER) (Durango, Torres-Silva, and Orozco-Duque 2023). In emergency medicine, NER and relation extraction (RE) have proven useful for identifying clinical concepts, events, and relationships.

Similarly, LLMs (Large Language Models) have also been applied for entity extraction from medical texts and text labeling, with the first studies demonstrating high reliability in the competition of this task, even in languages different from English (Akbasli, Birbilen, and Teksam 2025); in such studies, API (Application Programming Interface) requests to models deployed in external servers were implemented to complete this task, which implies that patient-data records are evaluated in the cloud.

However, many studies focus on highly represented languages such as English or Mandarin, with German being substantially underrepresented. German presents unique linguistic challenges, including flexible word order, compound structures and complex negation forms, which reduce model transferability and performance (Reinig and Markert 2023). Furthermore, in the case of LLM, the use of an API connected to external servers violates data protection rules in the hospital context.

Symptom extraction methods range from rule-based approaches to machine learning-assisted NER techniques (Durango, Torres-Silva, and Orozco-Duque 2023). However, few studies treat symptoms as the primary target entities (Luo et al. 2022). This is especially relevant for ED texts, which are written under time pressure and thus exhibit inconsistency and syntactic noise (Marshall et al. 2023). Improving model performance in this domain requires annotated corpora and real-world data integration.

Recent studies have shown that combining structured and unstructured data improves predictions of ED severity compared with using structured data alone (Bergsneider et al. 2024), emphasizing the value of anamnesis texts beyond administrative functions. Our approach builds upon this insight and uses fine-tuned NLP models to extract symptoms from German ED records—particularly from oncology patients, who often present with polysymptomatic profiles and high clinical complexity.

Unlike conventional approaches that search for predefined symptom lists (Bergsneider et al. 2024), we employ a holistic, data-driven method to map all affirmed symptoms directly from the narrative, reducing bias and preserving patient-centered documentation. Owing to their complex disease trajectories and the availability of high-quality registry data for retrospective validation, cancer patients serve as a testbed for this approach.

In this study, we first evaluate three NLP models—1) GLiNER, 2) Mistral-Nemo-Instruct-2407, and 3) a fine-tuned SCAI-BIO/BioGottBERT model—for their performance in symptom, negation, and anatomy recognition in German ED texts. We focus on their applicability in clinical practice, data protection compliance, and capacity to capture semantic nuance.

### 2 Methods

We analyzed emergency department data from the Klinikum Stuttgart, Germany, for patients aged 18 years and older from December 16, 2010, to September 12, 2024, totaling 256,453 ED visits. Simultaneously, the ONKOSTAR register, an SQL database for quality assurance and cancer registry reporting, contained 35,937 oncology patients within the same period.

Matching both datasets revealed 10,036 individuals with a cancer diagnosis and at least one ED visit, representing 3.9% of all ED patients. This result aligns with data from comparable emergency departments (Corlade-Andrei et al. 2025).

### Data preparation for nlp training

To fine-tune the machine learning models, we extracted 150 anamnesis texts from the emergency department database. All the texts were manually deidentified to allow annotation outside the hospital environment, ensuring compliance with data protection regulations and ethical standards. No metadata capable of identifying individual patients were stored or shared. Although the entire processing pipeline can be deployed on local hospital servers, manual deidentification serves as an additional safeguard to minimize any residual risk of patient reidentification.

Symptom expressions and relevant clinical features—such as preexisting conditions and documented procedures—were annotated manually by two independent annotators via Doccano, an open-source annotation tool for text data. In addition, we annotated anatomical references and negations. The inclusion of negations allowed us to differentiate affirmed from excluded symptoms (e.g., “no dyspnea”), whereas anatomical annotations helped localize symptoms within the body (e.g., “pain in the lower abdomen”).

Annotations were converted into token-level labels via the inside–outside–beginning (IOB) tagging scheme. Tokens at the start of a named entity were labeled ‘B-’, tokens inside an entity were labeled ‘I-’, and all other tokens were labeled ‘O’. This encoding enabled structured processing by the downstream NLP models.

The trained named entity recognition (NER) models were then used to extract structured information from the ED corpus. For each patient, we retained only the anonymized identifier along with basic clinical metadata (e.g., diagnosis, sex, age) and the extracted symptoms. All processing steps were executed within the hospital infrastructure, with strict adherence to data protection policies.

### NER Methods

NLP has proven effective in extracting insights from unstructured text data. Transformer-based models, a type of deep learning architecture, use self-attention mechanisms and parallel processing to analyze linguistic relationships and identify patterns in large datasets (Vaswani et al. 2023). These models consist of multiple layers that refine the data representation across different levels. By analyzing words in the context of entire sentences, they infer meaning and relationships more accurately. Parallel processing further enhances efficiency and speed (Vaswani et al. 2023).

Several studies have analyzed the performance of benchmark models applied to biomedical data, particularly in zero-shot settings^2^ (Abdul et al. 2024). However, all these models have been tested on clinical texts written in English. Furthermore, these implementations focus only on relevant clinical entities, whereas other relevant entities, such as negations, are not tested. For this reason, a more exhaustive analysis is required for NER models applied to German.

We utilize BERT-based models (bidirectional encoder representations from transformers, i.e., the ability to consider both the left and right contexts of words simultaneously^3^ (Devlin et al. 2019)), a widely adopted model specifically for text analysis (Devlin et al. 2019). Our previous research confirmed its suitability for capturing the semantic complexity of medical narratives (Diaz Ochoa et al. 2024). This transformer architecture is present in both the implemented models, the GlinNER model and the SCAI-BIO/BioGottBERT-based model.

Furthermore, we implemented Mistral-Nemo-Instruct-2407 as a zero-shot LLM model in a local deployment. We selected this model because of its high performance, simplicity in implementation, customization, and, more importantly, multilingual support^4^.

In summary, we evaluated the following three NLP models for automated symptom extraction from emergency department anamnesis texts:

- GLiNER (Generalized Language Model for Information Extraction and Recognition) Model^5^ ^6^ (Stepanov and Shtopko 2024) [Zero-shot]
- Mistral-Nemo-Instruct-2407 (Openweight transformer-based Large Language Model, LLM) Model^7^ (Jiang et al. 2023) [Zero-Shot]
- SCAI-BIO/BioGottBERT-based model (Scientific Curation & Annotation Initiative – Biomedical version/Biomedical GottBERT) Model^8^ (Diaz Ochoa et al. 2024) [Fine-tuning]

GliNER and Mistral-Nemo-Instruct-2407 were applied in a zero-shot setting, relying on their pretrained general language understanding without task-specific adaptation. In contrast, the SCAI-BIO/BioGottBERT model was further fine-tuned via a small-annotated dataset. Owing to its BERT-based architecture, it captures contextual dependencies and semantic nuances more effectively than traditional NLP methods such as bag-of-words or TF-IDF (term frequency-inverse document frequency) (Zhang et al. 2024).

This comparison enabled us to assess the reliability and applicability of zero-shot modeling vs. model fine-tuning for clinical information extraction. All the models were deployed locally on hospital servers, ensuring full compliance with data protection regulations and offering operational flexibility within clinical environments.

## Methods to increase the precision of extracted symptoms

For symptoms identified as “Schmerzen” (pain) that were not part of a compound term (e.g., “Kopfschmerzen” (headache)), we searched for anatomical references in the preceding or following entries. If such an anatomy was found, it was combined with the symptoms to provide a more specific description (e.g., anatomy + symptoms). This allowed us to include detailed pain symptoms, such as “Kopfschmerzen”, instead of the generic “Schmerzen” (see an example in Table 1).

**Table 1.** Example of original text including negated symptoms and symptoms detected by the model (original text is in German; in this table we deploy the corresponding English translation (E))

|  |  |  |
| --- | --- | --- |
| E | 4 weeks ago surgery for colon Ca with artificial exit on the right. In case of closure, resurgery with inpatient stay up to 2 weeks ago. For days significant Swelling of the right lower leg with redness and overheating. No dyspnea, no thoracic pain. | Swelling; redness; overheating |

<sup>4</sup> https://www.blog.brightcoding.dev/2025/02/24/ollama-mistral-revolutionizing-local-ai-deployment-with-open-source-power/ <sup>5</sup> https://github.com/urchade/GLiNER <sup>6</sup> https://huggingface.co/knowledgator/gliner-multitask-large-v0.5 <sup>7</sup> https://huggingface.co/Imstudio-community/Mistral-Nemo-Instruct-2407-GGUF <sup>8</sup> https://huggingface.co/SCAI-BIO/bio-gottbert-base/tree/main

Next, we cleaned the symptom entries by standardizing their format. All symptoms were converted to lowercase, with the first letter capitalized, and punctuation, such as periods and colons, was removed. For pain-related symptoms, the anatomy was directly combined with the term, so “head + ache” (“Kopf + Schmerzen”) became headache (“Kopfschmerzen”). To further streamline the data, we created a dictionary of common symptoms that included synonyms and word variations. This ensured that similar terms, such as “Verstopfung” and “Obstipation” (both obstipation), were standardized to a single term. We also accounted for word inflections (e.g., “husten” (cough) and “gehustet” (coughed)) to capture diverse forms of expression. Symptoms that matched the dictionary were replaced with the standardized term, whereas unmatched symptoms retained their original name.

## 3 Results

We developed and validated a fine-tuned NLP approach to extract affirmed symptoms from unstructured ED anamnesis texts. Oncology patients, a clinically complex and vulnerable group, were selected because of the availability of high-quality registry data, allowing robust retrospective validation. We transformed free text into structured symptom profiles and linked patients by symptom similarity, age group, and sex. This enabled the identification of clinically meaningful subgroups on the basis of symptom constellations.

### Annotation and evaluation strategy

The first step in developing our NLP model involved validating its ability to identify symptoms accurately via a structured annotation and evaluation workflow. Text annotation was carried out independently by two annotators. To assess the consistency of the annotations, interannotator agreement was measured. During the annotation process, we observed occasional differences between annotators, both in terms of the selected text spans and the assigned entity types (e.g., disease vs. symptom).

To account for such variations and evaluate model performance in a structured manner, we adopted four levels of evaluation granularity, following established standards in clinical named entity recognition (NER) (Tsai et al. 2006). These evaluation metrics range from a strict match—requiring exact agreement in both the text boundaries and the entity type—to more lenient metrics that tolerate partial overlaps or disregard the entity type. This allows for a more nuanced assessment of model performance (see Table 2).

**Table 2.** Different validation metrics implemented in this research.

| **Strict** | Exact match of both text boundaries and entity type |
| --- | --- |
| **Exact** | Exact match of text boundaries, regardless of entity type |
| **Partial** | Partial overlap of text boundaries, regardless of entity type |
| **Entity Type** | Matching entity type, allowing for some overlap with the gold standard annotation |

### Interannotator Agreement

To evaluate interannotator agreement, 20 narratives were annotated in parallel. While Cohen’s kappa is commonly used for document-level tasks, the F1 score is better suited for entity-level evaluations, such as symptom extraction. Table 3 shows the agreement across the four levels of granularity.

**Table 3.** Validation of interannotator agreement (n=20 narratives).

| **F1** | 0.72 | 0.58 | 0.68 | 0.56 |
| --- | --- | --- | --- | --- |
| **Precision** | 0.75 | 0.60 | 0.70 | 0.58 |
| **Recall** | 0.69 | 0.56 | 0.65 | 0.54 |

The highest agreement was observed for the entity type metric, with an F1 score of 0.72, precision of 0.75, and recall of 0.69. These metrics reflect how reliably symptoms were identified: precision measures how many of the extracted symptoms were correct, recall indicates how many of the relevant symptoms were captured, and the F1 score balances both measures into a single value (see Table 3).

The consistent performance across all the metrics confirmed the robustness of our annotation strategy. Using this result, small divergences in the annotators were corrected. With the redefined annotation guidelines, the other 130 examples were annotated for the final model fine-tuning.

### Model Comparison: Zero-Shot & LLM VS. Fine-Tuned Models

To identify the most suitable model for reliable symptom extraction, we compared zero-shot models— GLiNER and Mistral AI—with a fine-tuned transformer-based model (SCAI-BIO/BioGottBERT). For model validation, we used manual annotations of 150 emergency department narratives; 15% of the annotated narratives were used for model validation for all the tests.

### Performance of the GLiNER Model

The validation results for GLiNER are shown in Table 4. While the model performed reasonably well in recognizing symptoms (F1 score: 0.81), it showed substantial limitations in detecting negations (F1 score: 0.42; precision: 0.28), indicating a high false-positive rate.

**Table 4.** Validation results of the GliNER model (n=150 narratives).

|  |  |  |  |  |  |
| --- | --- | --- | --- | --- | --- |
| Total |  |  |  |  |  |
|  | F1 | 0.58 | 0.67 | 0.73 | 0.51 |
|  | Precision | 0.57 | 0.66 | 0.71 | 0.50 |
|  | Recall | 0.59 | 0.69 | 0.75 | 0.52 |
| Symptom |  |  |  |  |  |
|  | F1 | 0.81 | 0.69 | 0.76 | 0.67 |
|  | Precision | 0.77 | 0.66 | 0.73 | 0.64 |
|  | Recall | 0.85 | 0.72 | 0.80 | 0.70 |
| Negation |  |  |  |  |  |
|  | F1 | 0.42 | 0.39 | 0.40 | 0.39 |
|  | Precision | 0.28 | 0.26 | 0.27 | 0.26 |
|  | Recall | 0.81 | 0.75 | 0.78 | 0.75 |
| Anatomy |  |  |  |  |  |
|  | F1 | 0.56 | 0.62 | 0.70 | 0.53 |
|  | Precision | 0.64 | 0.71 | 0.80 | 0.60 |
|  | Recall | 0.5 | 0.55 | 0.65 | 0.47 |

### Performance of the Mistral AI Model

The Mistral-Nemo-Instruct-2407 model was evaluated locally (via local API access) and showed the weakest performance across all the validation categories (see Table 5). Symptom recognition achieved an F1 score of 0.46, and performance in negation detection decreased to an F1 score of 0.40, with particularly low precision values (e.g., 0.29 for negations).

**Table 5.** Validation results of the MISTRAL model (n=150 narratives).

|  |  |  |  |  |  |
| --- | --- | --- | --- | --- | --- |
| Total |  |  |  |  |  |
|  | F1 | 0.41 | 0.29 | 0.41 | 0.23 |
|  | Precision | 0.29 | 0.21 | 0.30 | 0.17 |
|  | Recall | 0.62 | 0.47 | 0.66 | 0.38 |
| Symptom |  |  |  |  |  |
|  | F1 | 0.46 | 0.31 | 0.40 | 0.30 |
|  | Precision | 0.32 | 0.21 | 0.28 | 0.21 |
|  | Recall | 0.83 | 0.55 | 0.73 | 0.53 |
| Negation |  |  |  |  |  |
|  | F1 | 0.40 | 0.066 | 0.27 | 0.052 |
|  | Precision | 0.29 | 0.048 | 0.20 | 0.03 |
|  | Recall | 0.65 | 0.10 | 0.44 | 0.08 |
| Anatomy |  |  |  |  |  |
|  | F1 | 0.29 | 0.19 | 0.29 | 0.16 |
|  | Precision | 0.21 | 0.14 | 0.21 | 0.11 |
|  | Recall | 0.48 | 0.32 | 0.49 | 0.27 |

### Performance of the SCAI-BIO/BioGottBERT base model

The fine-tuned SCAI-BIO/BioGottBERT base model significantly outperformed the GliNER model across all categories. It achieves the highest F1 score of 0.84 for symptom and negation recognition, demonstrating a strong balance between precision and recall. Particularly in negation handling, the model shows a substantial improvement, with an F1 score of 0.84, precision of 0.78, and recall of 0.90, making it notably more reliable in distinguishing between affirmed and negated symptoms.

**Table 6.**
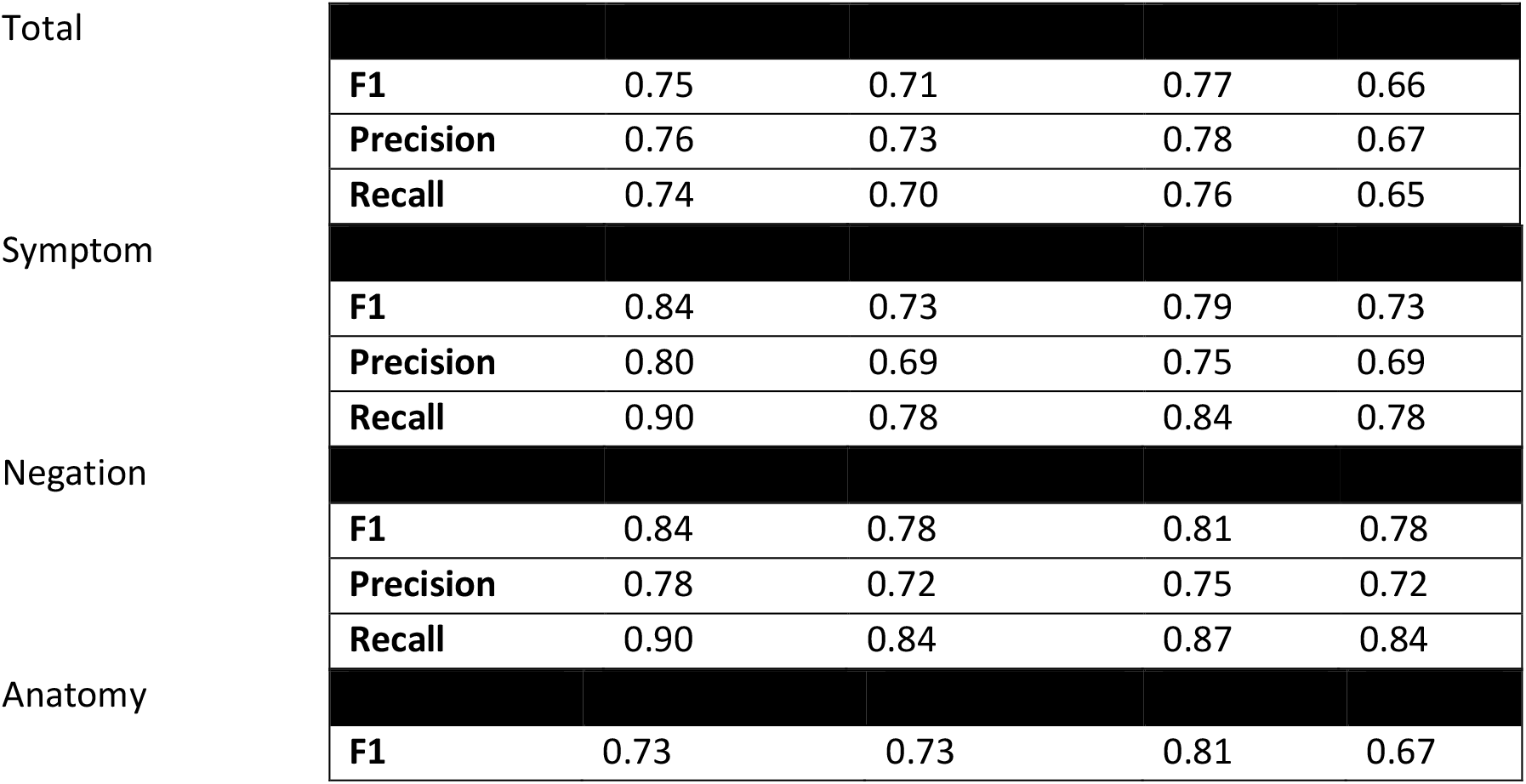
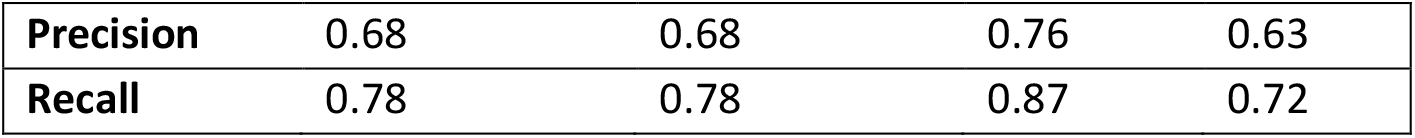
Validation results of the fine-tuned SCAI-BIO/BioGottBERT-based model.

|  |  |  |  |  |  |
| --- | --- | --- | --- | --- | --- |
| Total |  |  |  |  |  |
|  | F1 | 0.75 | 0.71 | 0.77 | 0.66 |
|  | Precision | 0.76 | 0.73 | 0.78 | 0.67 |
|  | Recall | 0.74 | 0.70 | 0.76 | 0.65 |
| Symptom |  |  |  |  |  |
|  | F1 | 0.84 | 0.73 | 0.79 | 0.73 |
|  | Precision | 0.80 | 0.69 | 0.75 | 0.69 |
|  | Recall | 0.90 | 0.78 | 0.84 | 0.78 |
| Negation |  |  |  |  |  |
|  | F1 | 0.84 | 0.78 | 0.81 | 0.78 |
|  | Precision | 0.78 | 0.72 | 0.75 | 0.72 |
|  | Recall | 0.90 | 0.84 | 0.87 | 0.84 |
| Anatomy |  |  |  |  |  |
|  | F1 | 0.73 | 0.73 | 0.81 | 0.67 |

|  |  |  |  |  |
| --- | --- | --- | --- | --- |
| **Precision** | 0.68 | 0.68 | 0.76 | 0.63 |
| **Recall** | 0.78 | 0.78 | 0.87 | 0.72 |

### Head-to-Head Evaluation of Model Performance

In summarizing model performance, we focused on the recognition of clinically relevant entities, particularly symptoms and negations. Since our pipeline excludes negated symptoms from further analysis, we refer to the final output as the SCAI-BIO/BioGottBERT-based-Filtneg model (SBBertFilNeg).

According to the comparative evaluation, GLiNER showed acceptable overall performance in symptom recognition but lacked reliability in detecting negations, limiting its clinical applicability. The Mistral AI model performed substantially worse across all the metrics, especially in recognizing contextual negations and anatomical references (see Figure 1). In contrast, the fine-tuned SBBertFilNeg model demonstrated consistently high scores across all entity types and validation levels, confirming its suitability for structured information extraction in emergency medicine.

**Figure 1.**
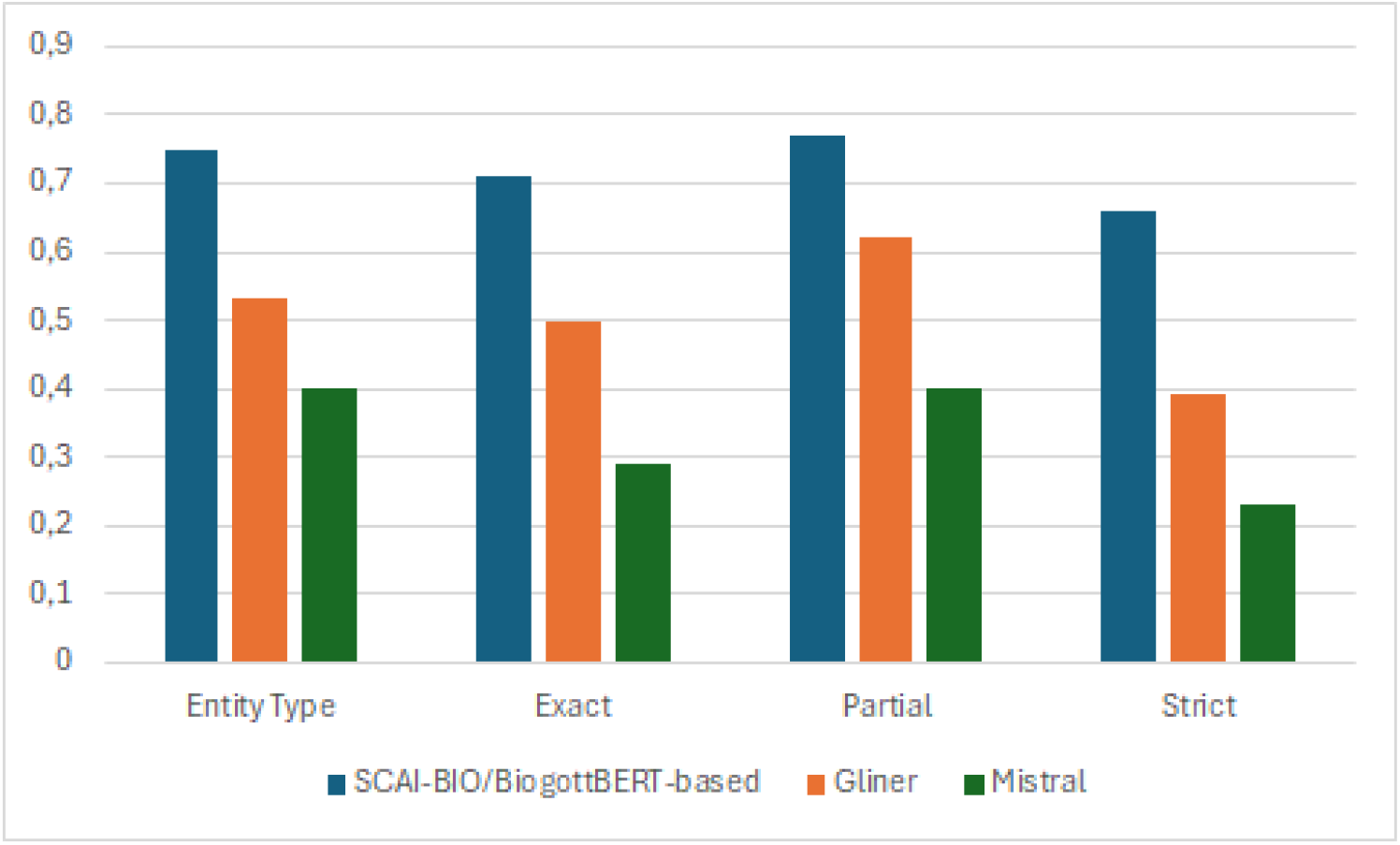
Comparison of the model validation results of the SCAI-BIO/BioGottBERT-based (blue), GLiNER (orange) and Mistral (gray) models considering the 4 different validation metrics (Table 2).

Thus, general zero-shot models can be implemented as a first proxy for NER tasks in a clinical context but are essentially incapable of capturing subtle variations in the language, particularly complex negation forms (which are often used in German).

In general, fine-tuned NLP models are more accurate in the recognition of entities in clinical texts, with a higher score with respect to zero-shot models (as visualized in Figure 1 for the four different kinds of validation methods) and are thus more appropriate for leveraging NER tasks in clinical narratives.

Finally, this result supports other studies analyzing benchmark models in biomedicine, where the LLM-Mistral has a lower performance than other benchmark models for NER tasks (for English, see Abdul et al. (Abdul et al. 2024)). Importantly, LLM models are implemented in an on-premises implementation, and such models can perhaps perform much better when implemented in a cloud implementation (Akbasli, Birbilen, and Teksam 2025).

There are two reasons that explain this low performance: local models generally have lower performance than online models accessed via API^9;^ furthermore, LLMs are designed to predict the next words or sentences that might be needed in finding entities in narratives. Thus, these results demonstrate that LLMs have different purposes and are very limited, especially when they are applied in the biomedical field. The limitations of these models in recognizing negations in the German language are also remarkable.

## 4 Discussion

### Context and Rationale of the Study

In emergency care, structured classification systems, such as ICD codes or the CEDIS, provide standardized snapshots of patient presentations. However, these predefined categories are inherently limited: they often fail to capture the nuanced, overlapping, and multisymptomatic nature of real-world clinical encounters.

Our approach instead captures the full spectrum of documented symptoms in an unbiased manner, directly from the physician’s free-text notes, without reducing them to predefined categories. This enables the identification of latent symptom constellations and subgroup-specific patterns, which may remain undetected in conventional documentation frameworks.

To achieve this goal, we implemented an NLP-based extraction pipeline for unstructured clinical narratives. Given the heterogeneous language, time pressure, and varying documentation styles in the ED, this task is nontrivial. Our study demonstrates how modern NLP techniques can overcome these challenges and enable fine-grained, data-driven symptom profiling, particularly in complex patient populations such as oncology patients.

### Methodological Foundation and State of the Art

Beyond textual heterogeneity and spelling errors, one key challenge lies in the diverse expression of negations, e.g., “no chest pain,” “patient denies chest pain,” or “chest pain not present,” which complicates symptom detection. This variation can greatly influence whether a symptom is interpreted as present or absent. Additionally, German and English differ substantially in grammar and lexical breadth. German’s flexible syntax and compound word structures pose significant obstacles for clinical NLP (Jantscher et al. 2023; Borchert et al. 2020).

Moreover, German medical texts in particular have been underrepresented in NLP model training, adding to the resource gap (Noll et al. 2023).

Nevertheless, as shown below, our study demonstrates that well-selected NLP techniques, specifically NER methods, are now capable of addressing such text nuances, with limited training effort.

### Model selection, BERT architecture and pretraining methodology

We compared three currently leading NLP model types with distinct architectures and learning strategies: for zero-shot learning, we implemented **GliNER** (Stepanov and Shtopko 2024) and **Mistral-Nemo-Instruct-2407** (Jiang et al. 2023); for fine-tuning, we implemented the **SCAI-BIO/BioGottBERT**: the fine-tuned BERT model, which was pretrained on German biomedical data (Diaz Ochoa et al. 2024).

BERT, as the underlying framework, was chosen for its capacity to integrate the bidirectional context and is crucial for capturing clinical semantics (Devlin et al. 2019) because of its strong performance in biomedical NLP tasks, including the automated coding of electronic health records (J. Wang et al. 2024).

Finally, a key requirement for all the models was deployability on local servers (on-premises configuration) to meet strict data protection standards.

## Results of the NLP models

Our findings demonstrate that zero-shot models offer a pragmatic entry point for clinical text analysis, especially in low-resource settings, and confirm previous results that demonstrate the capability of models from the BERT family for zero-shot learning (Y. Wang et al. 2023). However, even minimal fine-tuning with a small-annotated dataset led to marked performance improvements—particularly in negation handling. This highlights the efficiency of targeted fine-tuning and demonstrates that sustainable, locally applicable NLP pipelines are feasible without relying on large-scale generative AI models.

Furthermore, with respect to similar studies for data integration and modeling in EDs (Zhang et al. 2024), we have performed much more accurate symptom extraction, which allows fine-grained symptom analysis across the population. These results lay the foundation for data-driven risk stratification and hypothesis generation in oncology patients presenting to the ED—topics we address in a dedicated follow-up study.

### Patient-Centered Symptom Analysis Beyond Predefined Lists

A major strength of our approach lies in the fact that we do not rely on predefined symptom lists (Bergsneider et al. 2024). Instead, we allow symptoms to be recorded in free text, enabling patient-centered analysis. These unstructured symptom descriptions can be organized into cluster networks and presented in a structured way to clinicians, preserving the full diversity of individual patient expression. This approach captures not only the clinical presentation but also the physician’s interpretation, gender-related reporting differences, and sociocultural influences. While these aspects are inherently subjective and difficult to quantify, they are vital to understanding the true patient experience (Robinson 2023).

### Clinically Ready AI: Efficient, Local, and Privacy Compliant

Importantly, this analysis method was implemented with minimal computational resources, without reliance on large generative AI models. As such, the approach is well suited for local clinical infrastructures, adheres to strict data protection standards, and offers a sustainable, resource-efficient alternative to high-performance computing approaches. Our findings underscore that the future of digital healthcare may not lie in increasingly powerful AI systems but rather in intelligent, efficient modeling strategies that enhance clinical care while minimizing computational burden.

Comparative symptom frequency analysis revealed distinct clinical profiles in cancer patients compared with the general ED population, underscoring the importance of tailored symptom surveillance. The NLP-derived symptom profiles provide a solid basis for data-driven patient stratification and can be applied to enhance diagnose accuracy (Aissaoui Ferhi et al. 2024). The results of this study provide the basis for risk stratification and hypothesis generation in patients presenting to the Emergency Department, a topic of our upcoming follow-up study.

## 5 Conclusion

Symptoms recorded in unstructured ED notes contain critical clinical insights that extend beyond structured data fields such as CEDIS chief complaint systems. Our NLP pipeline captures all affirmed symptoms, preserving patient narrative complexity and individuality and enabling a patient-centered view of emergency care.

Fine-tuned, domain-specific models, particularly BERT-based approaches, outperform general LLMs and zero-shot methods in local model deployment. Even small, annotated datasets can significantly increase performance, making NLP feasible for real-world clinical use. Deployed entirely on local servers, our approach demonstrates that privacy-compliant, clinical-grade NLP is possible and implementable today.

This study bridges structured and unstructured data, enabling an enhanced layer of patient understanding. This study lays the foundation for symptom-based profiling, subgroup detection, and risk stratification in emergency oncology.

Moreover, it contributes to the vision of learning from cancer centers (LCCs), where clinical care, patient-centered research, and adaptive staff training are closely intertwined. Our work supports this model by transforming routine documentation into a dynamic, data-informed feedback system for continuous learning and quality improvement.

These results set the stage for hypothesis-driven stratification strategies in oncology patients presenting to the ED, which were further explored in a dedicated follow-up study.

## Data Availability

All data produced in the present study are available upon reasonable request to the authors

## 6 Ethical approval

This study was approved by the Ethics Committee at the Baden-Württemberg State Medical Association (Ethik-Kommissionder Landesärztekammer Baden-Württemberg), with approval number F-2024-105. The study was performed in compliance with the World Medical Association Declaration of Helsinki on Ethical Principles for Medical Research Involving Human Subjects and research regulations of the country. The informed consent requirement is waived owing to the state regulations of the Gesundheitsdatennutzungsgesetz (GDNG), owing to the retrospective, aggregated and anonymized nature of this study and of the database.

## 7 Contributions

JGDO, AK, TS, MK & NL discussed the first idea of applying NER for retrospective analysis of anamnesis texts; FEM & JGD developed and implemented the first prototypes; JM tested, implemented and validated all the NER models; JGDO & FEM implemented the models in the hospital; NL, TS & CM performed the data extraction; NL & JGDO performed the first evaluations of the NER; NL implemented the language inflections to improve the model outputs; JGDO implemented the clustering analysis; JGDO, NL, AK & MK designed the workflow; NL & JGDO performed the final evaluations as well as the workflow validation; NL, JGDO, TS & AK wrote the first draft of this study. JGDO, GI, MM & TS were responsible for the financial support of this project. NL, AK, MK & GI were responsible for the ethical approval of this study. All the authors contributed to the final version of this manuscript.

## 9 Acknowledgments

We want to thank Yi Wang (KI – Group, University Stuttgart) for very useful discussions on this topic. We also want to thank “Markus Krebs (Mainfranken, University Hospital Würzburg) for pointing out the relevance of this study for learning cancer centers.

## 11 Abbreviations

AI: Artificial Intelligence
API: Application Programming Interface
ED: Emergency Department
EHR: Electronic Health Record
LLM: Large Language Model
ML: Machine Learning
NER: Named Entity Recognition
NLP: Natural Language Processing
RE: Relation Extraction
TF-IDF: Term Frequency-Inverse Document Frequency

## FOOTNOTE

1 Currently working at NEC laboratories

2 See the NER dashboard at hugging face: https://huggingface.co/spaces/m42-health/clinical_ner_leaderboar

3 https://quickcreator.io/quthor_blog/bert-vs-large-language-models-differences/

4 https://www.blog.brightcoding.dev/2025/02/24/ollama-mistral-revolutionizing-local-ai-deployment-with-open-source-power/

5 https://github.com/urchade/GLiNER

6 https://huggingface.co/knowledgator/gliner-multitask-large-v0.5

7 https://huggingface.co/lmstudio-community/Mistral-Nemo-Instruct-2407-GGUF

8 https://huggingface.co/SCAI-BIO/bio-gottbert-base/tree/main

9 Using a synthetic dataset, we tested the performance of Mistral 7B in cloud computing and obtained an F1 score of 0.76; this indicates that AI implemented in the cloud is more effective than AI implemented locally.

## Bibliography

Abdul, Wadood M., Marco AF Pimentel, Muhammad Umar Salman, Tathagata Raha, Clément Christophe, Praveen K. Kanithi, Nasir Hayat, Ronnie Rajan, and Shadab Khan. 2024. “Named Clinical Entity Recognition Benchmark.” arXiv. 10.48550/arXiv.2410.05046.

Aissaoui Ferhi, Leila, Manel Ben Amar, Fethi Choubani, and Ridha Bouallegue. 2024. “Enhancing Diagnostic Accuracy in Symptom-Based Health Checkers: A Comprehensive Machine Learning Approach with Clinical Vignettes and Benchmarking.” Frontiers in Artificial Intelligence 7 (October):1397388. 10.3389/frai.2024.1397388.

Akbasli, Izzet Turkalp, Ahmet Ziya Birbilen, and Ozlem Teksam. 2025. “Leveraging Large Language Models to Mimic Domain Expert Labeling in Unstructured Text-Based Electronic Healthcare Records in Non-English Languages.” BMC Medical Informatics and Decision Making 25 (1): 1–9. 10.1186/s12911-025-02871-6.

Bergsneider, Brandon H., Terri S. Armstrong, Yvette P. Conley, Bruce Cooper, Marilyn Hammer, Jon D. Levine, Steven Paul, Christine Miaskowski, and Orieta Celiku. 2024. “Symptom Network Analysis and Unsupervised Clustering of Oncology Patients Identifies Drivers of Symptom Burden and Patient Subgroups With Distinct Symptom Patterns.” Cancer Medicine 13 (19): e70278. 10.1002/cam4.70278.

Borchert, Florian, Christina Lohr, Luise Modersohn, Thomas Langer, Markus Follmann, Jan Philipp Sachs, Udo Hahn, and Matthieu-P. Schapranow. 2020. “GGPONC: A Corpus of German Medical Text with Rich Metadata Based on Clinical Practice Guidelines.” arXiv. 10.48550/arXiv.2007.06400.

Corlade-Andrei, Mihaela, Radu-Alexandru Iacobescu, Viorica Popa, Alexandra Hauta, Paul Nedelea, Gabriela Grigorasi, Monica Puticiu, Roxana Elena Ciuntu, Andreea Ivona Sova, and Diana Cimpoesu. 2025. “Navigating Emergency Management of Cancer Patients: A Retrospective Study on First-Time, End-Stage, and Other Established Diagnoses in a High Turnover Emergency County Hospital.” Medicina 61 (1): 133. 10.3390/medicina61010133.

Devlin, Jacob, Ming-Wei Chang, Kenton Lee, and Kristina Toutanova. 2019. “BERT: Pre-Training of Deep Bidirectional Transformers for Language Understanding.” arXiv. 10.48550/arXiv.1810.04805.

Diaz Ochoa, Juan G., Faizan E. Mustafa, Felix Weil, Yi Wang, Kudret Kama, and Markus Knott. 2024. “The Aluminum Standard: Using Generative Artificial Intelligence Tools to Synthesize and Annotate Non-Structured Patient Data.” BMC Medical Informatics and Decision Making 24 (1): 409. 10.1186/s12911-024-02825-4.

Durango, María C., Ever A. Torres-Silva, and Andrés Orozco-Duque. 2023. “Named Entity Recognition in Electronic Health Records: A Methodological Review.” Healthcare Informatics Research 29 (4): 286–300. 10.4258/hir.2023.29.4.286.

Jantscher, Michael, Felix Gunzer, Roman Kern, Eva Hassler, Sebastian Tschauner, and Gernot Reishofer. 2023. “Information Extraction from German Radiological Reports for General Clinical Text and Language Understanding.” Scientific Reports 13 (1): 2353. 10.1038/s41598-023-29323-3.

Jiang, Albert Q., Alexandre Sablayrolles, Arthur Mensch, Chris Bamford, Devendra Singh Chaplot, Diego de las Casas, Florian Bressand, et al. 2023. “Mistral 7B.” arXiv. 10.48550/arXiv.2310.06825.

Kim, Jonathan, Anna Podlasek, Kie Shidara, Feng Liu, Ahmed Alaa, and Danilo Bernardo. 2025. “Limitations of Large Language Models in Clinical Problem-Solving Arising from Inflexible Reasoning.” arXiv. 10.48550/arXiv.2502.04381.

Luo, Xiao, Priyanka Gandhi, Susan Storey, and Kun Huang. 2022. “A Deep Language Model for Symptom Extraction From Clinical Text and Its Application to Extract COVID-19 Symptoms From Social Media.” IEEE Journal of Biomedical and Health Informatics 26 (4): 1737–48. 10.1109/JBHI.2021.3123192.

Marshall, Kyle, Ron Strony, Ben Hohmuth, and David K. Vawdrey. 2023. “New Coding Guidelines Reduce Emergency Department Note Bloat But More Work Is Needed.” Annals of Emergency Medicine 82 (6): 713–17. 10.1016/j.annemergmed.2023.07.023.

Munzone, Elisabetta, Antonio Marra, Federico Comotto, Lorenzo Guercio, Claudia Anna Sangalli, Martina Lo Cascio, Eleonora Pagan, et al. 2024. “Development and Validation of a Natural Language Processing Algorithm for Extracting Clinical and Pathological Features of Breast Cancer From Pathology Reports.” JCO Clinical Cancer Informatics, no. 8 (August), e2400034. 10.1200/CCI.24.00034.

Noll, Richard, Lena S. Frischen, Martin Boeker, Holger Storf, and Jannik Schaaf. 2023. “Machine Translation of Standardised Medical Terminology Using Natural Language Processing: A Scoping Review.” New Biotechnology 77 (November):120–29. 10.1016/j.nbt.2023.08.004.

Raghavan, Preethi, James L. Chen, Eric Fosler-Lussier, and Albert M. Lai. 2014. “How Essential Are Unstructured Clinical Narratives and Information Fusion to Clinical Trial Recruitment?” AMIA Summits on Translational Science Proceedings 2014 (April):218–23. https://www.ncbi.nlm.nih.gov/pmc/articles/PMC4333685/.

Reinig, Ines, and Katja Markert. 2023. “Can Current NLI Systems Handle German Word Order? Investigating Language Model Performance on a New German Challenge Set of Minimal Pairs.” arXiv. 10.48550/arXiv.2306.04523.

Robinson, William. 2023. “Epiphenomenalism.” In The Stanford Encyclopedia of Philosophy, edited by Edward N. Zalta and Uri Nodelman, Summer 2023. Metaphysics Research Lab, Stanford University. https://plato.stanford.edu/archives/sum2023/entries/epiphenomenalism/.

Stepanov, Ihor, and Mykhailo Shtopko. 2024. “GLiNER Multi-Task: Generalist Lightweight Model for Various Information Extraction Tasks.” arXiv. 10.48550/arXiv.2406.12925.

Tsai, Richard Tzong-Han, Shih-Hung Wu, Wen-Chi Chou, Yu-Chun Lin, Ding He, Jieh Hsiang, Ting-Yi Sung, and Wen-Lian Hsu. 2006. “Various Criteria in the Evaluation of Biomedical Named Entity Recognition.” BMC Bioinformatics 7 (1): 1–8. 10.1186/1471-2105-7-92.

Tybjerg, Karin. 2023. “Medical Anamnesis. Collecting and Recollecting the Past in Medicine.” Centaurus 65 (2): 235–59. 10.1484/J.CNT.5.135348.

Vaswani, Ashish, Noam Shazeer, Niki Parmar, Jakob Uszkoreit, Llion Jones, Aidan N. Gomez, Lukasz Kaiser, and Illia Polosukhin. 2023. “Attention Is All You Need.” arXiv. 10.48550/arXiv.1706.03762.

Wang, Jiajia, Jimmy Xiangji Huang, Xinhui Tu, Junmei Wang, Angela Jennifer Huang, Md Tahmid Rahman Laskar, and Amran Bhuiyan. 2024. “Utilizing BERT for Information Retrieval: Survey, Applications, Resources, and Challenges.” ACM Comput. Surv. 56 (7): 185:1-185:33. 10.1145/3648471.

Wang, Yue, Lijun Wu, Juntao Li, Xiaobo Liang, and Min Zhang. 2023. “Are the BERT Family Zero-Shot Learners? A Study on Their Potential and Limitations.” Artificial Intelligence 322 (September):103953. 10.1016/j.artint.2023.103953.

Zhang, Xingyu, Yanshan Wang, Yun Jiang, Charissa B. Pacella, and Wenbin Zhang. 2024. “Integrating Structured and Unstructured Data for Predicting Emergency Severity: An Association and Predictive Study Using Transformer-Based Natural Language Processing Models.” BMC Medical Informatics and Decision Making 24 (1): 1–13. 10.1186/s12911-024-02793-9.

